# Event-Wise Stability of Patient-Specific EEG-MEG Deep Learning Spike Detection in Clinical MEG

**DOI:** 10.64898/2026.08.18.26360638

**Authors:** Teppei Matsubara, Ryu Koda, Mark Richardson, Steven Stufflebeam

## Abstract

**Objective:** Computational magnetoencephalography (MEG) interictal epileptiform discharge (IED) detectors have mainly used generalized MEG-only models, whereas clinical MEG interpretation routinely integrates simultaneous electroencephalography (EEG) and includes MEG-unique or MEG-dominant discharges. We developed a patient-specific EEG-MEG IED detector and evaluated event-wise prediction stability across models and the effect of adding EEG to MEG-based prediction.

**Methods:** Seventeen patients undergoing clinical EEG-MEG evaluation for epilepsy were retrospectively analyzed. Clinically accepted dipole-review IEDs were treated as positive events, and nonannotated events were sampled as negatives. Logistic regression (LR), random forest (RF), and a lightweight three-dimensional ResNet were trained separately within each patient using EEG-only, MEG-only, and combined EEG-MEG (EMEG) inputs. Primary performance metrics were the area under the receiver operating characteristic curve (ROC-AUC) and average precision. Event-wise stability was assessed using rank disagreement, rank volatility, and class-aware distribution quotient analysis.

**Results:** Aggregate discrimination was high across models and modalities. Median ROC-AUCs for EEG, MEG, and EMEG were 0.850, 0.890, and 0.880 for LR; 0.880, 0.860, and 0.910 for RF; and 0.920, 0.960, and 0.960 for ResNet. Despite comparable aggregate performance, event-wise analysis revealed model-dependent prediction behavior. ResNet showed significantly lower non-IED rank volatility than classical machine learning models and lower non-IED rank disagreement, particularly compared with RF. Adding EEG to MEG was associated with more favorable class-aware event-wise positioning in most events, while MEG-unique/dominant cases showed greater relative MEG contribution.

**Conclusions:** Patient-specific EEG-MEG IED detection revealed clinically meaningful event-wise differences not captured by aggregate metrics. Simultaneous EEG complemented MEG-based detection, while MEG contribution remained prominent in MEG-dominant cases, supporting multimodal patient-specific IED event prioritization.

## 1. Introduction

MEG is a non-invasive neurophysiological tool that detects interictal epileptiform discharges (IEDs) with high spatiotemporal resolution (Rampp et al., 2019, Shiraishi et al., 2011, Tanaka and Stufflebeam, 2014). In clinical epilepsy practice, MEG is widely used to localize irritative zones through equivalent current dipole clustering of IEDs and contributes to presurgical evaluation and surgical planning, especially for targeting stereotactic electroencephalography (sEEG) implantation (Murakami et al., 2016). Compared with EEG, MEG signals are less affected by conductivity inhomogeneities of intervening tissues such as the skull and scalp (Hämäläinen et al., 1993), which is one reason MEG can provide more accurate source localization in selected clinical contexts.

Simultaneous surface EEG is routinely acquired during MEG recordings (Hari et al., 2018, Laohathai et al., 2021, Mosher and Funke, 2020) because EEG and MEG provide complementary information (Chikara et al., 2023, Tamilia et al., 2019), and EEG serves as a shared clinical reference among epileptologists and MEG reviewers. In clinical practice, MEG interpretation is fundamentally event-based: reviewers inspect individual candidate IEDs, evaluate their reproducibility, integrate EEG waveforms and MEG topography, and decide whether each event is suitable for dipole estimation. In addition, there are MEG-unique or MEG-dominant cases in which epileptiform abnormalities are more clearly identified on MEG than on EEG, highlighting the complementary but nonredundant nature of the two modalities (Ebersole and Wagner, 2018, Iwasaki et al., 2005). This discrepancy reflects several modality-specific factors, including differences in sensitivity to source orientation and configuration, sensor coverage, and signal detectability (Goldenholz et al., 2009, Matsubara et al., 2026).

Despite its clinical utility, MEG IED interpretation remains labor-intensive and highly dependent on expert visual review (Matsubara et al., 2020). Inter-reviewer variability remains substantial because IED morphology and spatial distribution vary across patients and across events even within the same patient. This event-wise variability makes definitive IED ground truth difficult to establish, particularly for subtle or ambiguous discharges.

Recent deep learning (DL)-based generalized MEG IED detectors have shown promising performance using large cross-patient cohorts (Fernandez-Martin et al., 2025, Hirano et al., 2022, Zheng et al., 2020). However, these approaches primarily relied on MEG signals alone and assumed shared IED patterns across patients, whereas clinical MEG interpretation incorporates simultaneous EEG and often depends on patient-specific IED morphology.

These limitations may be particularly relevant in MEG-dominant, MEG-unique, or low-confidence IEDs, where expert reviewers rely heavily on multimodal contextual interpretation rather than stereotyped IED morphology alone. We therefore developed a patient-specific IED detector using simultaneous EEG-MEG recording and compared DL with classical machine learning (ML) approaches. We hypothesized that DL would provide more stable event-wise prediction than classical ML and that adding EEG would improve MEG-based event positioning. We additionally examined whether MEG-unique/dominant cases showed greater relative MEG contribution, as a clinical test of modality complementarity.

## 2. Materials and Methods

### 2.1 Subjects

We retrospectively analyzed clinically acquired simultaneous EEG-MEG recordings from 17 patients who underwent routine clinical MEG evaluation for epilepsy at the Athinoula A. Martinos Center for Biomedical Imaging from November 2025 to May 2026 (Table 1). Patients were included if frequent IEDs were identified during routine clinical review and sufficient clinically accepted IED events were available for patient-specific model training, validation, and testing. This study was approved by Mass General Brigham’s institutional review board (#2010P001169).

**Table. 1.** Patient characteristics, interictal epileptiform discharge profiles, dipole distribution, and event counts.

|  | Subject | Age range at MEG | Etiology | MRI findings | IED morphology | Dipole distribution |  | IED type | Positive IED events |  |  |
| --- | --- | --- | --- | --- | --- | --- | --- | --- | --- | --- | --- |
|  |  |  |  |  |  | Lobar extent | Hemispheric extent |  | Train | Validation | Test |
| 1 | Subject 1 | 16–20 | PMG | Right parietal PMG | Spike-and-wave | 3 | 1 | Standard | 105 | 11 | 90 |
| 2 | Subject 3 | 31–35 | Cavernous malformation | Multiple cavernous malformations | Spike-and-wave | 2 | 2 | Standard | 42 | 11 | 29 |
| 3 | Subject 4 | 46–50 | Unknown | Nonlesional | Lateralized periodic discharge | 3 | 1 | Standard | 44 | 4 | 39 |
| 4 | Subject 5 | 16–20 | FCD | After left ATL (FCD type 2A) | Spike-and-wave | 3 | 2 | MEG dominant | 83 | 12 | 69 |
| 5 | Subject 6 | 46–50 | Abscess | Atrophy | Sharp-and-wave | 2 | 2 | Standard | 83 | 12 | 69 |
| 6 | Subject 7 | 41–45 | Unknown | Axon lesional | Spike-and-wave | 1 | 1 | MEG-unique | 84 | 7 | 74 |
| 7 | Subject 8 | 11–15 | Meningoencephalitis | After left occipital lobectomy | Spike-and-wave | 3 | 2 | MEG-dominant | 57 | 11 | 40 |
| 8 | Subject 9 | 21–25 | Stroke | Right parietal encephalomalacia | Spike-and-wave | 3 | 1 | Standard | 77 | 8 | 63 |
| 9 | Subject 10 | 36–40 | PMG | Right parietal PMG | Lateralized rhythmic delta activity | 3 | 2 | Standard | 63 | 9 | 51 |
| 10 | Subject 11 | 1–5 | TSC | Multiple tubers | Spike burst | 1 | 1 | Standard | 38 | 4 | 32 |
| 11 | Subject 12 | 6–10 | Unknown | Nonlesional | Polyspike-and-wave | 2 | 2 | Standard | 65 | 3 | 54 |
| 12 | Subject 13 | 11–15 | Ganglioglioma | After left ATL | Spike-and-wave | 2 | 2 | Standard | 53 | 5 | 46 |
| 13 | Subject 14 | 16–20 | Periventricular heterotopia | Bilateral periventricular heterotopia | Spike-and-wave | 3 | 1 | Standard | 75 | 9 | 66 |
| 14 | Subject 15 | 11–15 | Unknown | Nonlesional | Spike-and-wave | 1 | 1 | Standard | 34 | 4 | 26 |
| 15 | Subject 16 | 6–10 | Unknown | Nonlesional | Spike-and-wave | 2 | 1 | Standard | 87 | 6 | 78 |
| 16 | Subject 17 | 26–30 | Unknown | After left temporal arachnoid cyst resection | Spike-and-wave, paroxysmal fast | 2 | 1 | Standard | 81 | 8 | 71 |
| 17 | Subject 18 | 21–25 | Perinatal stroke | left MCA stroke | Sharp-and-wave | 1 | 1 | MEG-dominant | 20 | 9 | 9 |

Lobar extent indicates the number of lobes involved by clinically accepted dipoles; hemispheric extent indicates whether dipoles were localized to one or both hemispheres.
Abbreviations: ATL, anterior temporal lobectomy; FCD, focal cortical dysplasia; IED interictal epileptiform discharge; MCA, middle cerebral artery; MEG, magnetoencephalography; PMG, polymicrogyria; TSC, tuberous sclerosis complex

### 2.2 Clinical EEG-MEG recordings

Recordings included 306-channel whole-head MEG and simultaneous 70-channel scalp EEG using the 10-10 system, sampled at 2000 Hz (Matsubara et al., 2024, Tanaka et al., 2024). Each clinical study consisted of five raw recording files, typically 10 minutes each. MEG data were analyzed after standard MaxFilter processing with default clinical settings (Taulu et al., 2004), during which bad MEG sensors were interpolated.

Positive IED labels were derived from clinical MEG review. Specifically, IEDs were visually identified by expert review, T.M., a board-certified epileptologist, neurosurgeon, and clinical neurophysiologist, using simultaneous EEG for IED identification and MEG for dipole analysis. Events accepted for dipole estimation were treated as positive events. Therefore, positive labels reflected clinically selected dipole-review events rather than all visually suspected transient waveforms.

MEG-unique and MEG-dominant status was assigned during routine clinical EEG-MEG review based on visual inspection, not by a quantitative threshold. Cases were classified as MEG-unique when clinically accepted IEDs were identified on MEG without a recognizable scalp EEG correlate. MEG-dominant cases showed mixed patterns, in which some IEDs were visible on both EEG and MEG, whereas others were identifiable only or more clearly on MEG.

### 2.3 Preprocessing and epoch extraction

Continuous EEG-MEG data were processed in MNE-Python (Gramfort et al., 2013). EEG was re-referenced to the average reference. Data were band-pass filtered from 1 to 70 Hz and resampled to 200 Hz. Event-centered epochs were extracted from -100 ms to +150 ms relative to each annotated IED, yielding 250-ms windows. At 200 Hz, each epoch contained 50 temporal samples, which were used for subsequent model input. Bad EEG channels were excluded on a patient-specific basis before feature extraction and model training.

### 2.4 Positive and negative event sampling

Positive samples were clinically accepted IED events. Negative samples were randomly selected from nonannotated time points within the same raw recordings. To reduce contamination by adjacent IEDs, candidate negative samples within 1.0 s of positive events were excluded. Negative samples were generated at a 3:1 negative-to-positive ratio.

### 2.5 Patient-specific train-validation-test design

All models were trained separately within each patient. Testing was separated from model development at the raw-file level rather than by random epoch splitting. Within each patient, held-out raw recordings were reserved exclusively for testing, and no epochs from these recordings were used for training, validation, or model selection. Training and validation samples were drawn only from the remaining development recordings.

Within the train recordings, validation data were selected using spike-aware 20-s temporal blocks with a 10-s buffer removed from the surrounding training data to reduce temporal leakage. Block selection prioritized mixed IED/non-IED content and targeted at least 3 IEDs, approximately 10% of training IEDs, and approximately 15% of valid training duration when possible. Because validation data were selected in contiguous blocks, the final number of validation IEDs could exceed the target in some patients. For ResNet models, validation blocks were used for epoch selection. For LR and RF models, fixed hyperparameters were used, and validation scores were computed for reference but were not used to tune model parameters. The final test recordings were not used during model selection. No offline data augmentation was applied.

Three modality conditions were evaluated: EEG-only, MEG-only, and combined EEG-MEG, hereafter referred to as EEG, MEG and EMEG.

### 2.6 Classical ML models

Following the three modality conditions above, classical ML models were trained using EEG, MEG, and EMEG inputs. Hand-extracted features were constructed to summarize amplitude- and topography-related information from each event. This feature-based design also allowed us to assess the relative contribution of amplitude and topographic information, as well as the relative contribution of EEG and MEG signals in EMEG models.

Two classical ML models were trained: logistic regression (LR) (Hosmer et al., 2013) and random forest (RF) (Breiman, 2001). LR used feature standardization, balanced class weights, and a maximum of 1000 iterations. RF used 300 trees, unrestricted depth, minimum split size of 4, minimum leaf size of 2, balanced class weights, and parallel training.

Classical features were grouped into four categories: EEG amplitude, EEG topography, MEG amplitude, and MEG topography. EEG amplitude features summarized channel-wise amplitude and temporal morphology, including peak-to-peak amplitude, absolute maximum, line length, slope, and global field power. EEG topographic features summarized peak-time channel distributions, including raw and z-normalized topographies and spatial laterality/anterior-posterior summaries.

MEG amplitude features were computed separately for gradiometers and magnetometers and included peak-to-peak amplitude, absolute maximum, line length, and global field power. MEG topographic features were also computed separately for gradiometers and magnetometers using peak-time sensor amplitude distributions and their z-normalized topographies. Because topographic features were sensor-wise representations, they formed a larger feature group than amplitude-derived summary features.

For EMEG models, feature contribution was assessed by grouping model-derived feature importance into EEG and MEG components. For LR, contribution was based on absolute standardized model coefficients. For RF, contribution was based on feature importance values. The MEG-to-EEG contribution ratio (M/E ratio) was defined as the summed MEG contribution divided by the summed EEG contribution within the EMEG model to represent the relative contribution of MEG versus EEG features.

### 2.7 3D spatiotemporal topographic representation

For DL, each epoch was converted into a 3D spatiotemporal volume. At each time point, sensor amplitudes were projected onto fixed two-dimensional topographic grids according to the sensor layout. EEG was represented on a 16 × 16 grid. MEG gradiometers and magnetometers were represented separately on 32 × 32 grids. The 50 temporal frames were stacked to create a volume with dimensions time × height × width (3D).

For EEG models, the EEG volume was used as a single input. For MEG models, gradiometer and magnetometer volumes were processed as two separate branches. For EMEG models, EEG, gradiometer, and magnetometer volumes were processed as three separate branches and fused at the embedding level.

### 2.8 3D ResNet architecture

The DL model was a lightweight three-stage 3D ResNet (Hara et al., 2018, He et al., 2016) implemented in PyTorch. Each input branch began with a 3D convolution using a temporal-spatial kernel of 7 × 3 × 3 and 32 output channels, followed by batch normalization and ReLU activation.

The ResNet backbone consisted of three residual stages. Stage 1 used two residual blocks without downsampling and preserved 32 feature channels. Stage 2 contained two residual blocks; the first block downsampled the feature maps with a stride of 2 × 2 × 2 and increased the number of feature channels from 32 to 64. Stage 3 also contained two residual blocks; the first block again downsampled the feature maps with a stride of 2 × 2 × 2 and increased the number of feature channels from 64 to 128. Each residual block contained two 3D convolutions with 3 × 3 × 3 kernels, batch normalization, ReLU activation, and either an identity shortcut or a 1 × 1 × 1 projection shortcut when dimensions changed.

For EEG models, the output of the ResNet backbone was summarized by adaptive global 3D average pooling and passed to a fully connected head with dropout, a 64-unit hidden layer, ReLU activation, dropout, and a final single-logit output.

For MEG and EMEG models, the same ResNet backbone was used as a modality-specific feature extractor. Each branch produced a 128-dimensional embedding after adaptive global 3D average pooling, dropout, and a linear projection. In MEG models, gradiometer and magnetometer embeddings were concatenated and passed to a classifier with a 128-unit hidden layer, ReLU activation, dropout, and a final single-logit output. In EMEG models, EEG, gradiometer, and magnetometer embeddings were concatenated and passed through the same classifier structure.

Models were trained for up to 30 epochs using batch size 8, learning rate 1 × 10^−5^, Adam optimization, weight decay 1 × 10^−4^, and binary cross-entropy with logits. The loss function was weighted according to the negative-to-positive class ratio (i.e., 3). Model selection was performed using the validation set.

### 2.9 Performance evaluation and group analysis

Primary performance metrics were the area under the receiver operating characteristic curve (ROC-AUC) and average precision (AP). Accuracy, precision, recall, and F1 were calculated internally but were not treated as primary outcomes because they depend on threshold selection, whereas our primary goal was to evaluate model ranking performance and event-wise score patterns.

For group-level analysis, subject-level metrics were summarized using the median and interquartile range (IQR). Subject-level paired comparisons were performed using two-sided Wilcoxon signed-rank tests. Holm correction was applied within each analysis family, and Holm-adjusted p values < 0.05 were considered statistically significant. ROC-AUC and AP were compared across LR, RF and ResNet.

Modality contribution was assessed for EMEG models using the M/E ratio defined in Section 2.6.

### 2.10 Event-wise analysis

Because aggregate ROC-AUC and AP can remain similar even when individual event predictions differ substantially, event-wise analyses were performed to assess prediction stability across modalities and models.

For each test event, prediction scores from EEG, MEG, and EMEG were compared within each model. Because raw prediction-score distributions differed across models and modalities, direct comparison of raw scores was not considered sufficient. Therefore, event-wise prediction scores were converted to normalized within-model ranks after aligning the direction of favorability; higher raw scores were considered more favorable for IED events, whereas lower raw scores were considered favorable for non-IED events. After this alignment, higher ranks indicated more favorable prediction for both event classes.

Rank disagreement was defined as the absolute difference between the EEG and MEG ranks for each event. This metric quantified how differently EEG and MEG prioritized the same IED or non-IED event.

Rank volatility was defined as the range of ranks across EEG, MEG, and EMEG modalities for each event. This metric quantified the stability of event-wise prediction ordering across modalities.

Finally, class-aware distribution quotient analysis (DQ) was performed to evaluate how adding EEG altered MEG-based event-wise prediction. For clarity, subject and model indices are omitted below; all quantities were computed separately within each subject and model. Let *s_i_*_,*k*_ be the prediction score for event *i* under modality condition *k*, where *k* ∈ {*EEG*, *MEG*, *EMEG*}, and let *y_i_* ∈ {0, 1} denote the event label.

For each modality condition *k*, *μ*_0,*k*_ and *μ*_1,*k*_ were defined as the median prediction scores of non-IED and IED events, respectively, and *IQR*_0,*k*_ and *IQR*_1,*k*_ were defined as the corresponding IQRs. The midpoint between the two class distributions was defined as

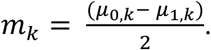

The scale term was defined as

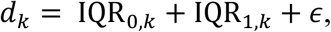

where *ε* is a small positive constant added as a scale floor for numerical stability. The class-aware normalized margin was then defined as

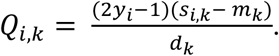

With this definition, higher *Q_i_*_,*k*_ values indicate more favorable positioning for both event classes: higher scores for IED events and lower scores for non-IED events. The event-wise EMEG gain over MEG was defined as

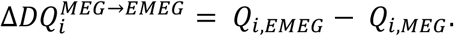

A positive Δ*DQ^MEG^*^→*EMEG*^ indicates that EMEG positioned the event more favorably than MEG alone relative to the class-specific score distributions. Finally, the fraction of events with Δ*DQ^MEG^*^→*EMEG*^ > 0 was calculated across all test events, representing the proportion of events for which adding EEG improved class-aware score positioning relative to MEG alone.

Two EEG-only ResNet models, from subjects 11 and 18, showed non-informative outputs, with nearly identical scores across IED and non-IED events. These subjects were retained in the aggregate ROC-AUC and AP summaries in Fig. 1A where applicable, but were excluded entirely from subsequent event-wise rank and DQ analyses in Fig. 2 to keep the analyzed subject set consistent across models and modalities.

**Figure 1.**
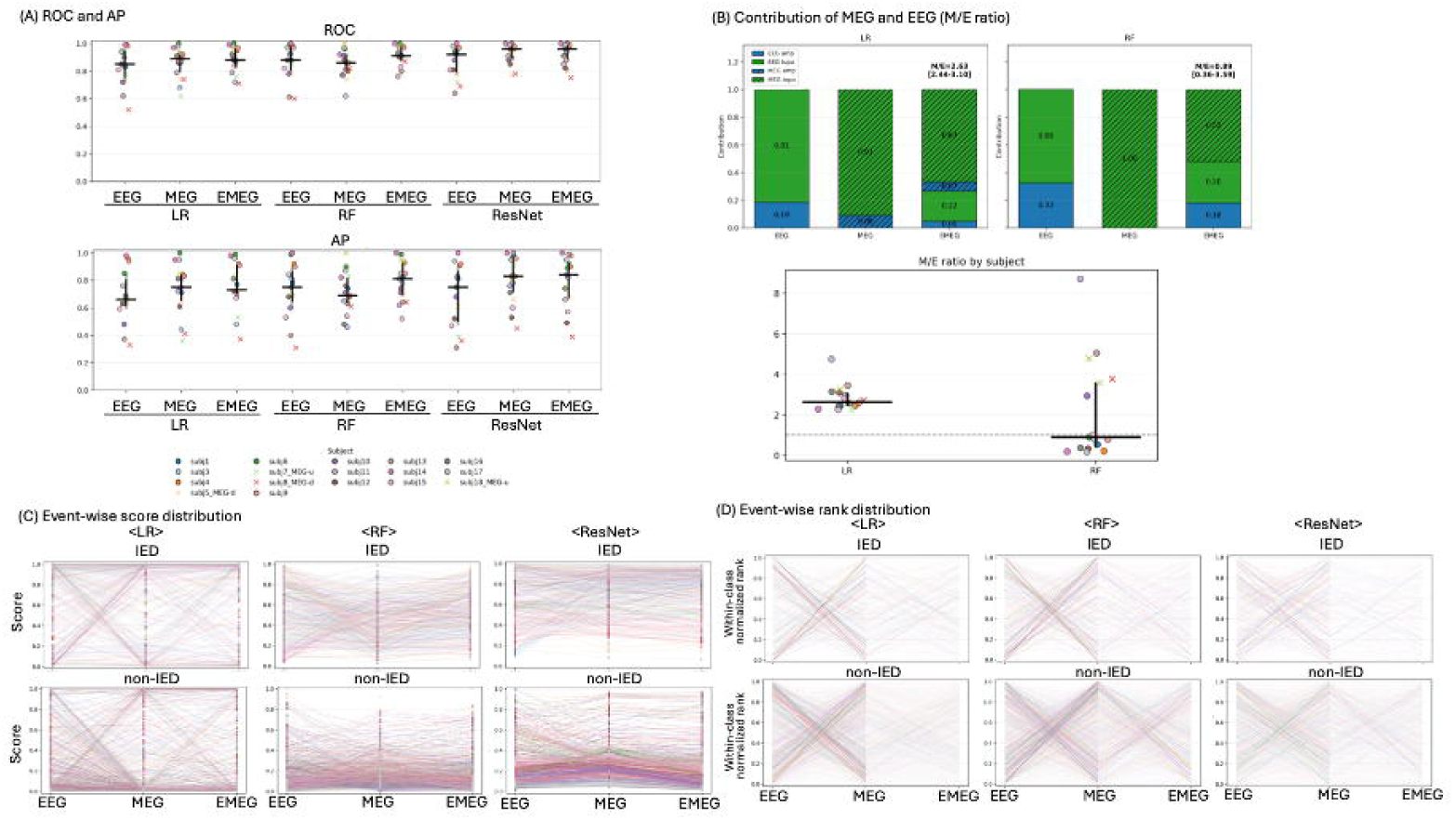
Summary results across all patients (A) The area under the receiver operating characteristic curve (upper panel) and average precision (lower panel) across models and modality conditions. Subject-level data points were overlaid with group medians and interquartile ranges (IQRs) in black and 95% confidence intervals in gray. Subject colors are consistent between Figures 1 and 2. MEG-dominant or MEG-unique cases are marked with crosses.(B) Classical machine learning feature contributions. Stacked bar plots show feature contributions for logistic regression and random forest across modality conditions (EEG, MEG and EMEG). Hand-extracted features were grouped into four categories: EEG amplitude, EEG topography, MEG amplitude and MEG topography. The lower panel shows the subject-level MEG-to-EEG contribution ratio (M/E ratio) in EMEG models. (C) Event-wise score distributions for all subjects, shown separately for interictal epileptiform discharge (IED) events (upper panels) and non-IED events (lower panels). Each line represents the trajectory of an individual event across modality conditions. (D) Event-wise rank distributions across all subjects, shown separately for IED events (upper panels) and non-IED events (lower panels). Event-wise prediction trajectories are shown using original-direction within-class normalized ranks. Ranks closer to 1 correspond to higher predicted IED scores, whereas ranks closer to 0 correspond to lower predicted IED scores within each event class. Thus, favorable prediction corresponds to ranks closer to 1 for IED events and closer to 0 for non-IED events. Dense overlapping lines indicate concentrated event-wise prediction trajectories across modality conditions.

**Figure 2.**
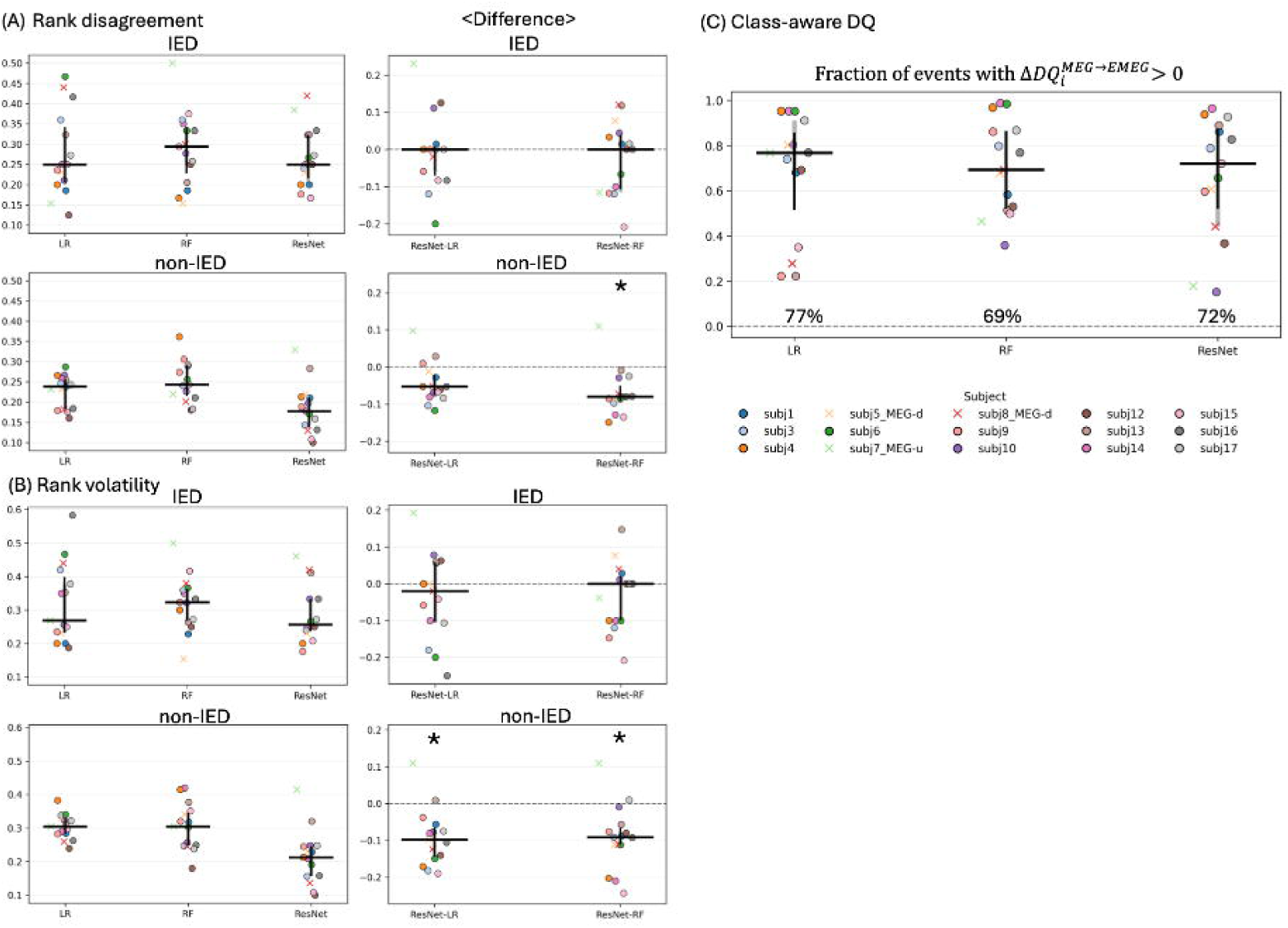
Subject-level statistics for event-wise analysis (A) Rank disagreement. Left panels show EEG-MEG rank disagreement for each model and subject. Lower values indicate less disagreement between EEG and MEG event ordering. Right panels show paired subject-level differences in rank disagreement between ResNet and classical machine learning models. (B) Rank volatility. Left panels show rank volatility across EEG, MEG, and EMEG for each model and subject. Lower values indicate more stable event-wise ordering across modality conditions. Right panels show paired subject-level differences in rank volatility between ResNet and classical machine learning models. (C) Class-aware distribution quotient (DQ) analysis. Fraction of events with positive ΔDQ from MEG to EMEG is shown for each model, where positive ΔDQ indicates that EMEG provided more favorable class-aware event positioning than MEG alone. Asterisks indicate differences that remained statistically significant after Holm correction (Holm-adjusted p < 0.05).

## 3. Results

### 3.1 Overall model performance was high across modalities and models

Overall discrimination performance was high across models and modality conditions, indicating that patient-specific IED detection achieved near-ceiling aggregate performance in many subjects (Fig. 1A upper). For ROC-AUC, median values were 0.850 [IQR, 0.750–0.940] for EEG, 0.890 [0.790–0.920] for MEG, and 0.880 [0.830–0.960] for EMEG using LR. Corresponding values were 0.880 [0.800–0.970], 0.860 [0.800–0.920], and 0.910 [0.870–0.980] using RF, and 0.920 [0.790–0.960], 0.960 [0.890–0.980], and 0.960 [0.880–0.990] using ResNet.

AP showed a similar pattern. Median AP values (Fig. 1A lower) were 0.660 [0.610–0.805] for EEG, 0.750 [0.650–0.840] for MEG, and 0.730 [0.700–0.910] for EMEG using LR; 0.750 [0.640–0.870], 0.690 [0.630–0.820], and 0.810 [0.690–0.930] using RF; and 0.750 [0.495–0.870], 0.830 [0.710–0.960], and 0.840 [0.670–0.930] using ResNet.

Paired model comparisons showed significantly higher MEG ROC-AUC for ResNet than for LR and RF. Median paired differences were 0.040 [0.010–0.080] for ResNet minus LR (Holm-adjusted p = 0.025) and 0.080 [0.020–0.130] for ResNet minus RF (p = 0.035). For AP, the largest ResNet advantage was also observed in MEG, particularly against RF, with a median paired difference of 0.110 [0.060–0.230], although this difference did not remain significant after Holm correction (p = 0.40). In contrast, paired differences for EEG and EMEG were smaller and had IQRs overlapping zero.

Thus, aggregate ROC-AUC and AP indicated strong overall detector performance but did not fully capture how individual events were prioritized across modalities or models.

### 3.2 Classical ML feature contributions showed complementary EEG and MEG information

Feature contribution analysis of the classical ML models was used to examine which types of hand-extracted features supported IED prediction across modality conditions (Fig. 1B). Contributions were first grouped by feature type, namely amplitude- and topography-related features within EEG and MEG, and then summarized in EMEG models as the relative M/E ratio.

Across both LR and RF, topographic features contributed more prominently than amplitude features (Fig. 1B upper). This pattern was particularly evident in MEG models, where MEG topographic features dominated MEG amplitude features, suggesting that the spatial distribution of sensor-level activity was more informative than amplitude summary measures alone. A similar tendency was also observed in EEG models, although the relative balance between amplitude and topography varied across subjects and model types.

In EMEG models, both EEG and MEG features contributed to prediction, supporting a complementary rather than redundant role of the two modalities. The balance between EEG and MEG differed between LR and RF. In LR, MEG features contributed more strongly than EEG features in all subjects, with a median M/E ratio of 2.630 [2.440–3.100]; all 17 subjects had an M/E ratio greater than 1 (Fig. 1B lower), indicating greater MEG than EEG contribution. In RF, the median M/E ratio was 0.890 [0.360–3.590], with 8 of 17 subjects showing greater MEG than EEG contribution. Notably, these 8 subjects included all 4 clinically identified MEG-unique or MEG-dominant cases, indicating that RF captured greater relative MEG dependence in cases where epileptiform abnormalities were clinically more apparent on MEG than on EEG. Thus, LR showed a consistently MEG-weighted pattern, whereas RF showed more variable, patient-specific weighting between EEG and MEG.

### 3.3 Event-wise analysis revealed model-dependent stability not captured by ROC/AP

Because aggregate ROC-AUC and AP were high across models, we next examined event-wise prediction behavior. Event-wise score and rank trajectories showed that models with similar aggregate discrimination could differ substantially in how they ordered individual IED and non-IED events. In representative trajectory patterns (Fig. 1C and in representative cases in Figs. 3C, D and 4C, D), LR tended to show broader, vertically dispersed event distributions, RF showed intermediate dispersion, and ResNet showed a more compact event-wise distribution, suggesting greater stability of event ordering.

**Figure 3.**
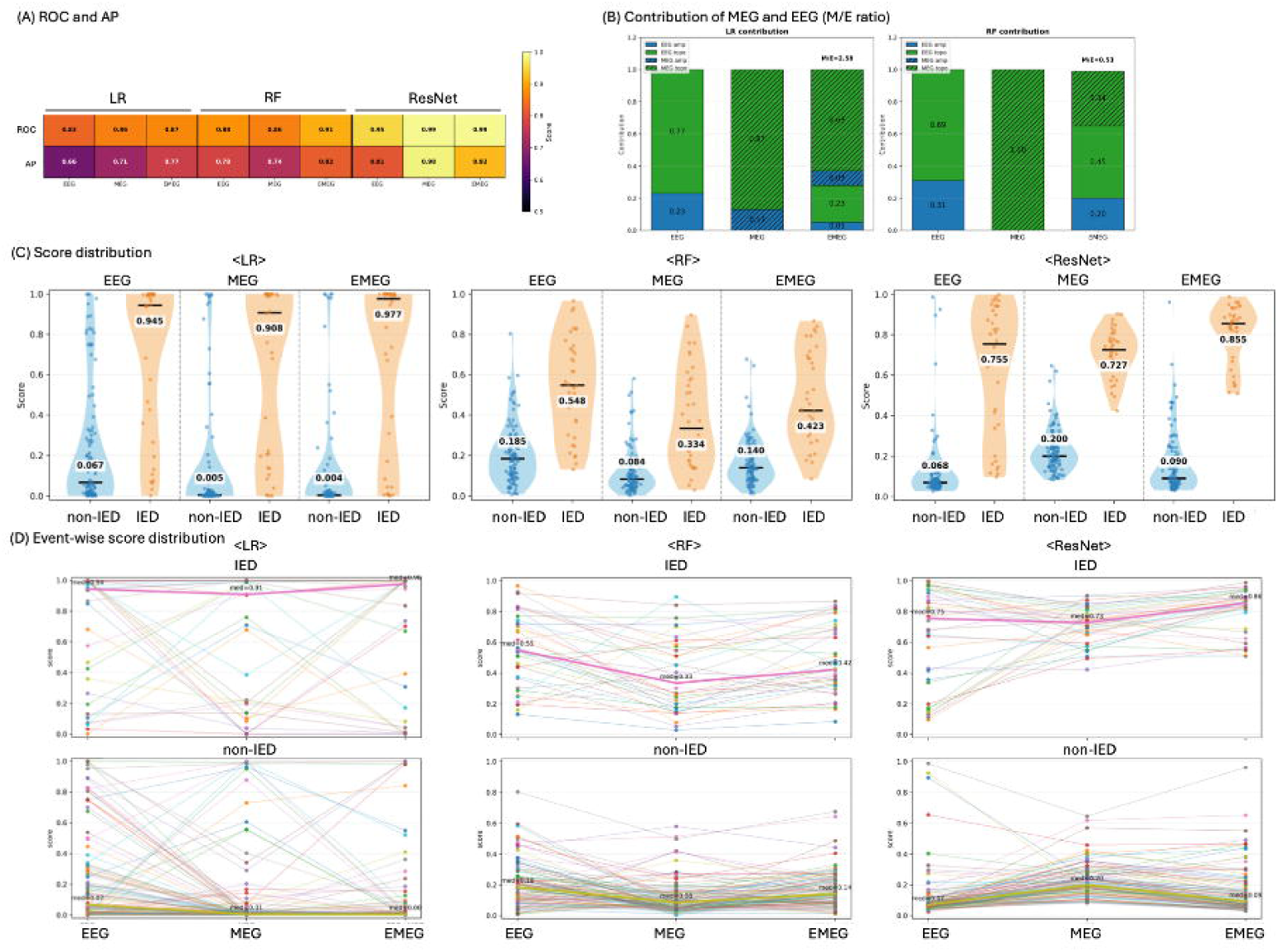
Representative subject (subject 1) (A) The area under the receiver operating characteristic curve and average precision across models and modality conditions. Aggregate performance was generally high and comparable across modalities and models. (B) Relative modality contributions in logistic regression (LR) and random forest (RF). Each stacked bar represents amplitude and topographic feature contributions for each modality condition. (C) Score distributions for each model and modality, shown separately for non-interictal epileptiform discharge (IED) events (blue) and IED events (orange). (D) Event-wise score distributions for IED events (upper panels) and non-IED events (lower panels). Event-wise scores showed marked cross-modality variability in LR and RF, whereas ResNet demonstrated more stable prediction trajectories across modalities.

This visual impression was further summarized using rank distribution (Fig. 1D) and quantified using rank disagreement and rank volatility. For non-IED events, median EEG-MEG rank disagreement (Fig. 2A left panels) was 0.238 [0.183–0.256] for LR, 0.243 [0.215–0.291] for RF, and 0.177 [0.137–0.212] for ResNet. For IED events, corresponding values were 0.250 [0.206–0.342], 0.294 [0.228–0.342], and 0.250 [0.215–0.323].

Rank volatility across EEG, MEG, and EMEG showed a similar pattern (Fig. 2B left panels). For non-IED events, median volatility was 0.305 [0.284–0.327] for LR, 0.305 [0.248–0.346] for RF, and 0.213 [0.157–0.246] for ResNet. For IED events, median volatility was 0.269 [0.233–0.399] for LR, 0.324 [0.269–0.363] for RF, and 0.257 [0.235–0.333] for ResNet.

Paired comparisons confirmed greater non-IED event-wise stability for ResNet than for classical ML models (Figs. 2A, B right panels). For non-IED events, ResNet minus LR showed a median rank disagreement difference of -0.053 [-0.075 to -0.020] (p = 0.075), and ResNet minus RF showed -0.080 [-0.091 to -0.050] (p = 0.026). For non-IED rank volatility, ResNet minus LR was -0.098 [-0.144 to -0.065] (p = 0.021), and ResNet minus RF was -0.091 [-0.111 to -0.066] (p = 0.023). In contrast, IED-event differences were smaller and did not remain significant after Holm correction.

Class-aware DQ analysis was then used to evaluate whether adding EEG to MEG shifted event-wise predictions in a favorable direction. Median ΔDQ from MEG to EMEG was positive for all models: 0.053 [-0.001–0.592] for LR, 0.052 [0.015–0.448] for RF, and 0.077 [0.003–0.237] for ResNet. The fraction of events with positive ΔDQ was 0.768 [0.515–0.858] for LR, 0.692 [0.522–0.864] for RF, and 0.720 [0.520–0.875] for ResNet (Fig. 2C). These findings suggest that adding EEG to MEG was associated with more favorable class-aware event-wise score positioning for a majority of test events, supporting the complementary role of EEG during IED prediction.

### 3.4 Representative cases illustrate clinically interpretable event-wise behavior

Two representative cases were examined to illustrate how event-wise model behavior differed despite high aggregate performance. The first representative case showed generally good ROC-AUC and AP across models and modalities (Fig. 3A), but event-wise score distributions differed markedly across algorithms (Fig. 3C and D). LR showed a broader and more vertically dispersed distribution of event scores, whereas ResNet produced a more compact event-wise distribution, consistent with the cohort-level rank-volatility analysis (Fig. 1C, D).

The MEG-unique case further illustrated the clinical relevance of multimodal interpretation (Fig. 4). In this case, EEG-only prediction was more challenging, whereas MEG and EMEG models better captured clinically selected IED events. Feature contribution analysis also showed greater relative MEG contribution in the MEG-unique case than in the representative standard case (Fig. 4B versus Fig. 3B), including an RF M/E ratio of 3.59. This finding was consistent with the clinical observation that IEDs were more clearly expressed in MEG than in EEG. These representative examples demonstrate that aggregate ROC-AUC and AP alone are insufficient to characterize clinically relevant model behavior, and that event-wise trajectories provide additional insight into modality complementarity and model stability.

**Figure 4.**
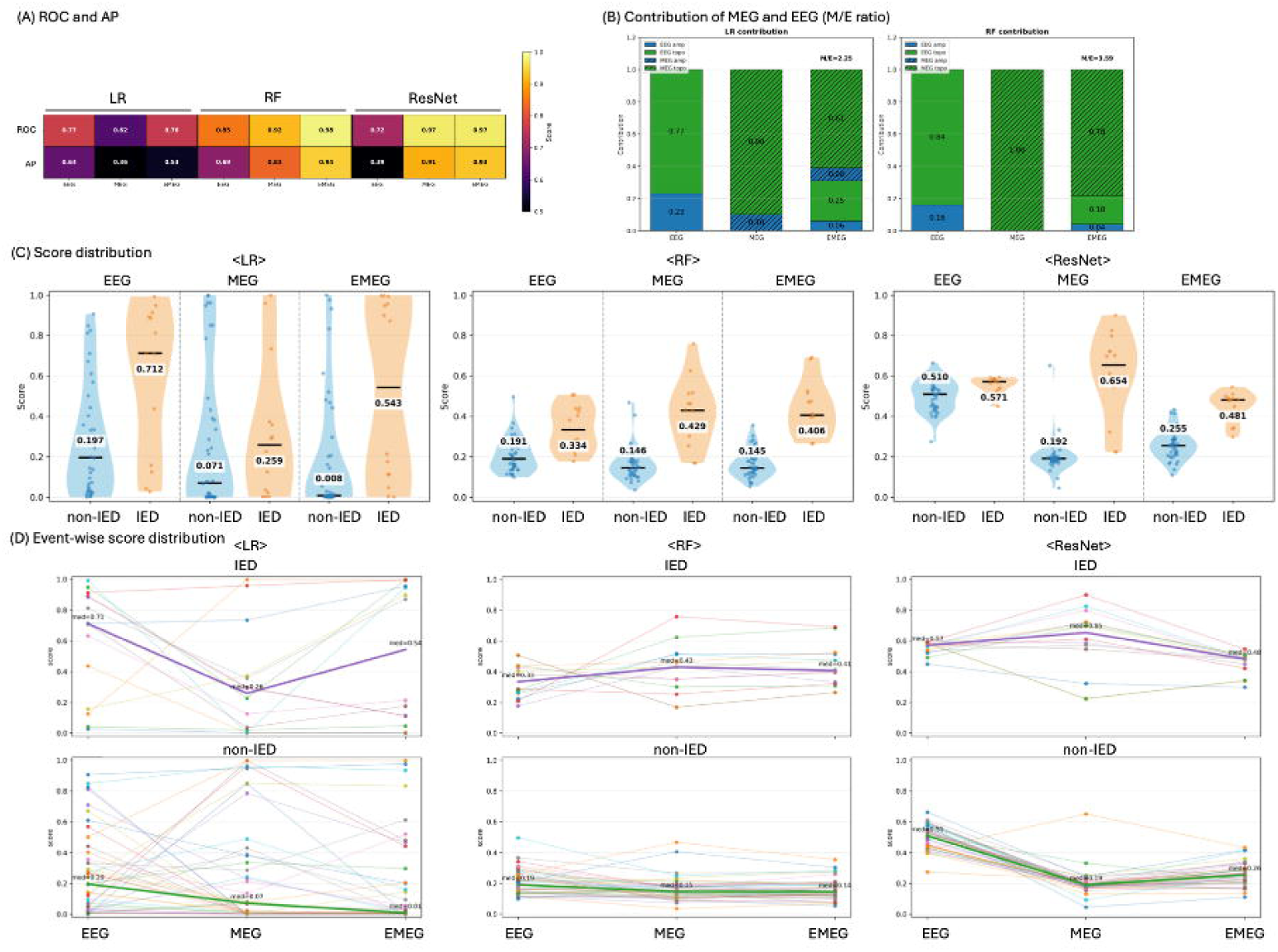
Representative MEG-unique subject (subject 7) Panels are arranged as in Figure 3. In this MEG-unique case, the M/E contribution ratio in random forest was higher than that in the representative standard subject (3.59 versus 0.53), indicating greater relative MEG contribution to EMEG prediction. EEG-only models showed weaker class separation, with generally lower area under the receiver operating characteristic curve values and greater overlap between interictal epileptiform discharge (IED) and non-IED score distributions, particularly in random forest and ResNet. This pattern was consistent with the clinical observation that IEDs were more clearly expressed in MEG than in EEG.

## 4. Discussion

In this study, we developed a patient-specific IED detector using simultaneous EEG-MEG recordings acquired during routine clinical MEG evaluation. First, all models achieved high aggregate discrimination performance (Fig. 1A), suggesting that patient-specific IED prediction is feasible using clinically derived labels. Second, despite similar ROC-AUC and AP values across models and modalities, event-wise analyses revealed substantial differences in prediction behavior (Fig. 1C, D). In particular, paired analyses confirmed greater non-IED event-wise stability for ResNet than for classical ML models (Fig. 2A, B). Third, adding EEG to MEG was associated with more favorable class-aware event-wise score positioning in the majority of test events (Fig. 2C). These findings support patient-specific DL-assisted review as a potentially useful approach for MEG interpretation, not only by detecting IED events but also by improving the stability of event prioritization across modalities.

### 4.1 Relationship to generalized MEG IED detectors

ML- and DL-based IED detection has been studied extensively in EEG (da Silva Lourenco et al., 2021, Nhu et al., 2022), and recent studies have expanded this framework to MEG using DL-based approaches. Earlier MEG studies demonstrated that DL can learn clinically meaningful features from MEG recordings and achieve high performance for IED detection. EMS-Net used a Multiview DL architecture to detect MEG IEDs from raw MEG data, combining single-channel and multichannel representations (Zheng et al., 2020). FAMED further extended MEG IED detection toward automated dipole analysis by using a segmentation-based DL framework trained on a large clinical cohort, demonstrating high classification performance and clinically plausible dipole localization (Hirano et al., 2022). STIED also emphasized the importance of spatiotemporal MEG features by combining temporal waveform and spatial topographic information in the supervised DL framework (Fernandez-Martin et al., 2025).

These studies established the feasibility of generalized MEG-based IED detection. However, most were trained across patients and primarily used MEG signals alone. In contrast, routine clinical MEG interpretation is inherently patient-specific and incorporates simultaneous EEG: reviewers compare candidate events within the same patient, assess reproducibility, and determine whether events are suitable for dipole analysis. Therefore, our approach should be viewed as complementary to generalized MEG IED detectors rather than as a replacement. Whereas generalized detectors may provide broad first-pass screening, patient-specific EEG-MEG models may refine event prioritization based on the patient’s own IED morphology and modality profile.

### 4.2 Complementary roles of EEG and MEG in clinical IED prediction

Beyond this methodological distinction from prior generalized MEG detectors, our results support the complementary roles of EEG and MEG in patient-specific IED prediction. In clinical MEG interpretation, EEG provides a shared clinical reference for epileptologists and MEG reviewers, whereas MEG provides dense sensor coverage and accurate source-localization information (Hari et al., 2018, Laohathai et al., 2021, Mosher et al., 2020). The two modalities are therefore complementary rather than redundant.

In classical ML models, both EEG and MEG features contributed to EMEG prediction (Fig. 1B). Topographic features contributed more strongly than amplitude features, suggesting that the spatial pattern of sensor-level activity carried important information for IED classification. The M/E ratio further showed that MEG contribution was substantial, particularly in LR, while RF demonstrated more variable patient-specific weighting between EEG and MEG. This variability is clinically plausible, because relative usefulness of EEG and MEG differs across patients depending on IED source orientation, depth, spatial extent, background activity, and sensor-level detectability (Matsubara et al., 2026).

DQ analysis provided an event-wise assessment of how adding EEG altered MEG-based prediction (Fig. 2C). Adding EEG to MEG shifted class-aware event positioning in a favorable direction for a majority of test events across models. This does not imply that EEG is always superior or should dominate MEG-based prediction. Rather, EEG provided additional event-level information that was associated with more favorable model ranking for many IED and non-IED events, consistent with routine clinical practice in which EEG and MEG are interpreted together (Laohathai et al., 2021).

### 4.3 MEG-unique and MEG-dominant cases

The importance of modality complementarity is especially evident in MEG-unique and MEG-dominant cases. In these patients, epileptiform abnormalities are visible only on MEG or are more clearly expressed on MEG than on scalp EEG (Ebersole et al., 2018, Iwasaki et al., 2005). Such dissociation is expected from known modality-specific differences, including sensitivity to source orientation, spatial sampling, signal-to-noise ratio, and distortion by intervening tissues (Ahlfors et al., 2010, Hunold et al., 2016). Therefore, an algorithm that treats EEG and MEG as interchangeable inputs may fail to reflect the actual clinical value of MEG in these cases.

The representative MEG-unique case illustrated this point (Fig. 4). EEG-only prediction was relatively challenging, whereas MEG and EMEG better captured clinically selected IED events. In addition, RF feature contribution analysis showed greater relative MEG contribution in the MEG-unique case, consistent with the clinical observation that IEDs were more clearly expressed in MEG. At the subject level (Fig. 1B), MEG-unique and MEG-dominant cases tended to show higher M/E ratios, again supporting greater MEG dependency in these cases.

The difference between LR and RF contribution patterns should be interpreted cautiously. M/E ratios are model-dependent indicators of feature use rather than direct physiological measures. Nevertheless, the finding that MEG-unique and MEG-dominant cases showed greater relative MEG contribution, particularly in RF, supports the clinical relevance of patient-specific modality weighting.

Clinically, these findings suggest that patient-specific models may be particularly useful in MEG-dominant or MEG-unique situations. In such cases, the goal is not simply to reproduce EEG-visible IEDs, but to assist reviewers in identifying and prioritizing MEG-relevant IEDs that may be subtle, spatially focal, or poorly represented on scalp EEG.

### 4.4. Importance of event-wise analysis beyond ROC-AUC and AP

A key finding of this study is that aggregate ROC-AUC and AP did not fully capture clinically relevant model behavior. In patient-specific IED detection, high aggregate performance can coexist with substantial event-wise instability (Fig. 1C, D): the same IED event may be assigned a high score in one modality and a low score in another, or may be prioritized differently by LR, RF, and ResNet. Such instability matters clinically because MEG review is event-based: reviewers inspect individual candidate discharges, assess reproducibility, and decide whether they should be used for dipole estimation.

Rank disagreement and rank volatility quantified this event-wise stability. ResNet showed lower rank disagreement and rank volatility than classical ML models, especially for non-IED events (Fig. 2A, B). This indicates that the 3D spatiotemporal representation learned by ResNet provided more stable non-IED event prioritization than hand-extracted features. This is clinically relevant because false-positive candidate events increase reviewer workload and reduce confidence in AI-assisted review.

The DQ analysis further showed that adding EEG to MEG shifted class-aware event positioning in most test events. Conceptually, DQ is a Cohen’s d-like standardized margin measure that normalizes each event’s class-aware score position by within-class dispersion, rather than a formal inferential statistic. Thus, simultaneous EEG contributed useful event-level information even when MEG-only aggregate performance was high. These findings support event-wise analysis as an important evaluation strategy for clinical AI tools intended to support expert neurophysiological review.

### 4.5 Clinical implications

The proposed framework is best understood as an assistive tool for clinical MEG review. It is not intended to replace expert interpretation or automated ECD analysis. Instead, a patient-specific detector could help prioritize candidate events in subsequent recordings from the same patient, reduce reviewer burden, and improve consistency of event selection for dipole analysis. This may be particularly useful in patients with frequent IEDs, variable IED morphology, MEG-dominant findings, or ambiguous low-confidence events.

Another potential advantage of the patient-specific approach is that it aligns with how clinical MEG review is performed. Expert reviewers do not evaluate each event in isolation using a universal rule; rather, they learn the patient’s characteristic IED morphology, compare events across the recording, and integrate EEG, MEG topography, and source localization. A patient-specific model can approximate this within-patient learning process. The present results suggest that DL, by learning 3D spatiotemporal topographic patterns, may provide a more stable representation of this patient-specific event structure than classical ML models based on hand-extracted features.

### 4.6 Limitations

This study has several limitations. First, the sample size was modest, and the analysis was retrospective and conducted at a single center using a single EEG-MEG system. Although the patient-specific design reduces the need for a large cross-patient training cohort, larger datasets will be needed to assess robustness across epilepsy types, age groups, recording conditions, MEG hardware, and EEG montages. Second, positive labels were derived from clinical MEG review by a single expert reviewer and reflected events accepted for dipole estimation. Therefore, inter-rater reliability could not be assessed. Although these clinically selected labels are relevant to the intended clinical workflow, they do not represent all visually suspicious transients or all true epileptiform events. Thus, the model was trained to reproduce clinically selected dipole-review events rather than a definitive biological ground truth.

Third, negative samples were selected from nonannotated time points. Although temporal exclusion around positive IEDs was used to reduce contamination, some negative samples may have included unmarked subtle IEDs or ambiguous transients. This limitation is inherent to clinical IED detection, where exhaustive ground truth is difficult to establish.

### 4.7 Future directions

Future studies should evaluate patient-specific EEG-MEG IED detection prospectively within the clinical MEG workflow, focusing on reviewer time, false-positive burden, consistency of dipole-review event selection, and agreement with expert reviewers. It will also be important to test whether a model trained on earlier recordings can prioritize IED candidates in later recordings from the same patient. Finally, patient-specific and generalized approaches should be compared in the same dataset, as a hybrid framework may combine broad candidate screening with patient-specific event prioritization based on each patient’s IED morphology and modality profile.

### 4.8 Conclusions

Patient-specific IED detection using simultaneous EEG-MEG achieved high aggregate performance and revealed clinically meaningful event-wise differences across models and modalities. ResNet showed significantly lower non-IED rank volatility than classical ML models and lower non-IED rank disagreement, particularly compared with RF. Adding EEG to MEG was associated with more favorable class-aware event positioning in the majority of test events. These findings support the complementary role of EEG in clinical MEG interpretation and suggest that patient-specific DL-assisted review may improve the stability and clinical usability of IED event prioritization.

## Data Availability

All data produced in the present study are available upon reasonable request to the authors.

## Abbreviations

AP: average precision
DL: deep learning
DQ: distribution quotient (DQ)
EEG: electroencephalography
IED: interictal epileptiform discharge
IQR: interquartile range
LR: logistic regression
MEG: magnetoencephalography
ML: machine learning
RF: random forest
ROC-AUC: area under the receiver operating characteristic curve

